# Unboxing unhealthy food marketing: An analysis of the extent and nature of unhealthy food marketing to children on YouTube in Canada using the CHOPPED artificial intelligence system

**DOI:** 10.64898/2026.11.12.26367461

**Authors:** Sara Al Sharani, Tyler Williamson, Spencer Fietz, Leticia Radin Pereira, Charlene Elliott, Monique Potvin Kent, Inara Lalani, Sadeeb Hossain, Zahra Shakeri Hossein Abad, Shelly Yin, Rachel Prowse, Dana Lee Olstad

## Abstract

**Background:** Unhealthy food marketing negatively affects children’s dietary patterns. However, little is known about the extent and nature of unhealthy food marketing to children on YouTube due to reliance on manual assessment methods.

**Objectives:** To use a novel artificial intelligence (AI) system (CHild Oriented food Product Promotion Evaluation and Detection system, CHOPPED) to examine the extent and nature of unhealthy food marketing in YouTube videos suggested to Avatar accounts created for children and adults based on age and gender.

**Methods:** Over two months of continuous monitoring, CHOPPED collected marketing instances played before, during, and after videos suggested by YouTube’s algorithm to six child and two adult accounts that were not signed in and differed by age (2-4, 5-8, 9-12, ≥18 years) and gender across four Canadian cities (Calgary, Vancouver, Toronto, Halifax). CHOPPED characterized the extent and nature of monthly child-targeted unhealthy food marketing instances during 24-hour and typical 2-hour daily viewing periods.

**Results:** CHOPPED collected 60,633 marketing instances. No food marketing was shown to child accounts 2-4 years. On average, child accounts (5-12 years) were shown 25 monthly unhealthy food marketing instances during a 2-hour daily viewing period, 24 of which were child-targeted. The boy’s account (9-12 years) was shown a significantly higher monthly frequency of child-targeted unhealthy food marketing instances (340, 70.2%) than the boy’s account (5-8 years; 241, 66.2%), the girl’s account (5-8 years; 261, 60.6%), the man’s account (224, 57.7%), and the woman’s account (263, 54.7%) during a 24-hour (p<0.05) but not a 2-hour daily viewing period. Monthly frequencies of some core child-targeted techniques shown to the boy’s account (9-12 years) were also higher than other accounts during a 24-hour (p<0.05) but not a 2-hour daily viewing period.

**Conclusions:** This study pioneered AI-driven analysis of the extent and nature of unhealthy food marketing to children on YouTube. Children’s potential exposure to such marketing on YouTube may be low. There was some evidence that the older boys’ account was shown more monthly child-targeted unhealthy food marketing instances and of some core child-targeted techniques than other accounts. On YouTube, food marketing may target a general audience rather than children by age and gender.

## BACKGROUND

Dietary patterns develop in childhood and can persist into adulthood [1]. Globally, children’s dietary patterns are concerning, with more than half of children consuming poor quality diets [2]. In Canada, children’s dietary patterns are also poor, with average scores of just 40 out of 80 on the Healthy Eating Food Index-2019 [3]. Additionally, children’s diets are high in ultra-processed foods and low in fruits and vegetables [4,5]. A key environmental factor that shapes children’s dietary patterns is the marketing of unhealthy foods and beverages (hereafter “food”), such as those high in sodium, sugar, and/or saturated fats [6]. Unhealthy food marketing influences children’s dietary patterns and body weight by influencing their product awareness and preferences, and subsequently their food requests, food purchases, and consumption [6].

Food companies are increasingly shifting their marketing spending towards digital marketing, where the influence on children’s food preferences, attitudes, and consumption may be greater than on non-digital media [7,8]. YouTube, the most popular online video sharing platform among children [9–11], has been shown to drive nearly three times more product purchases than other platforms [10].

The extent of unhealthy food marketing to children on digital media refers to the frequency of unhealthy food marketing they may encounter [12]. A study of YouTube’s most popular child-targeted videos found that foods were marketed more frequently than any other product within these videos (38.0% of marketing instances), and a majority of these marketing instances promoted unhealthy foods (56.3%) [13]. Another study in Canada showed that children were exposed to 1.96 food marketing instances in 30 minutes of watching YouTube videos, with 87.0% promoting unhealthy foods [14]. The nature of food marketing, which refers to the persuasive marketing techniques used to target children, plays a key role in increasing sales of marketed foods [12]. One study that examined energy drink marketing to children on YouTube, Facebook, and Twitter found that the most employed marketing techniques were thrill-seeking themes, presence of celebrities, and product health and personal benefit claims [15].

A key limitation of prior studies is that they all used manual methods to assess the extent and nature of unhealthy food marketing to children on YouTube and other digital media. Although manual methods can provide detailed assessments of marketing content and persuasive techniques, they are labour intensive, highly subjective and limit the volume of digital content that can feasibly be analyzed [16]. This is particularly the case for digital media due to the persistent and highly targeted nature of digital marketing.

Artificial intelligence (AI), which refers to computer systems capable of executing tasks typically associated with human intelligence [17], can help to analyze large volumes of marketing by automating difficult, time-consuming, or sometimes unattainable tasks for researchers. We recently developed a novel AI system capable of monitoring the extent and nature of food marketing on digital media. The CHild Oriented food Product Promotion Evaluation and Detection system (CHOPPED) consists of seven modules and uses commercially available computing services and custom-made machine learning models. These seven modules include marketing data collection, feature extraction, food and brand identification, healthfulness classification, marketing technique classification, policy adherence classification, and aggregation and visualization. An initial prototype of the AI system [18] performed poorly, and thus, it was abandoned and a new system—CHOPPED—was developed by a new team co-led by DLO and TW. This new system was used in the present study. A forthcoming publication describes development and validation of the CHOPPED AI system in further detail (Al Sharani et al, unpublished observations).

The aim of this study was to use our newly developed AI system (CHOPPED) to examine the extent and nature of unhealthy food marketing in YouTube videos suggested to Avatar accounts created for children and adults based on age and gender.

## METHODS

### Study Design

This study collected and analysed marketing instances from YouTube using our AI system, CHOPPED. It used publicly available data and did not involve human participants; therefore, it was deemed exempt from ethical review by the Conjoint Health Research Ethics Board at the University of Calgary.

### Data Collection

In order to collect marketing instances from YouTube’s recommended videos, one boy and one girl Avatar account were created for each of three age categories including preschool (2-4 years), younger (5-8 years), and older (9-12 years) children informed by the Advertising Standards Authority’s previous work in the UK (Table 1) [19]. We also created two adult accounts for one woman and one man (≥18 years) to assess whether there were differences in the extent and nature of food marketing instances shown to child and adult accounts. The process of data collection is described in Figure 1.

**Figure 1.**
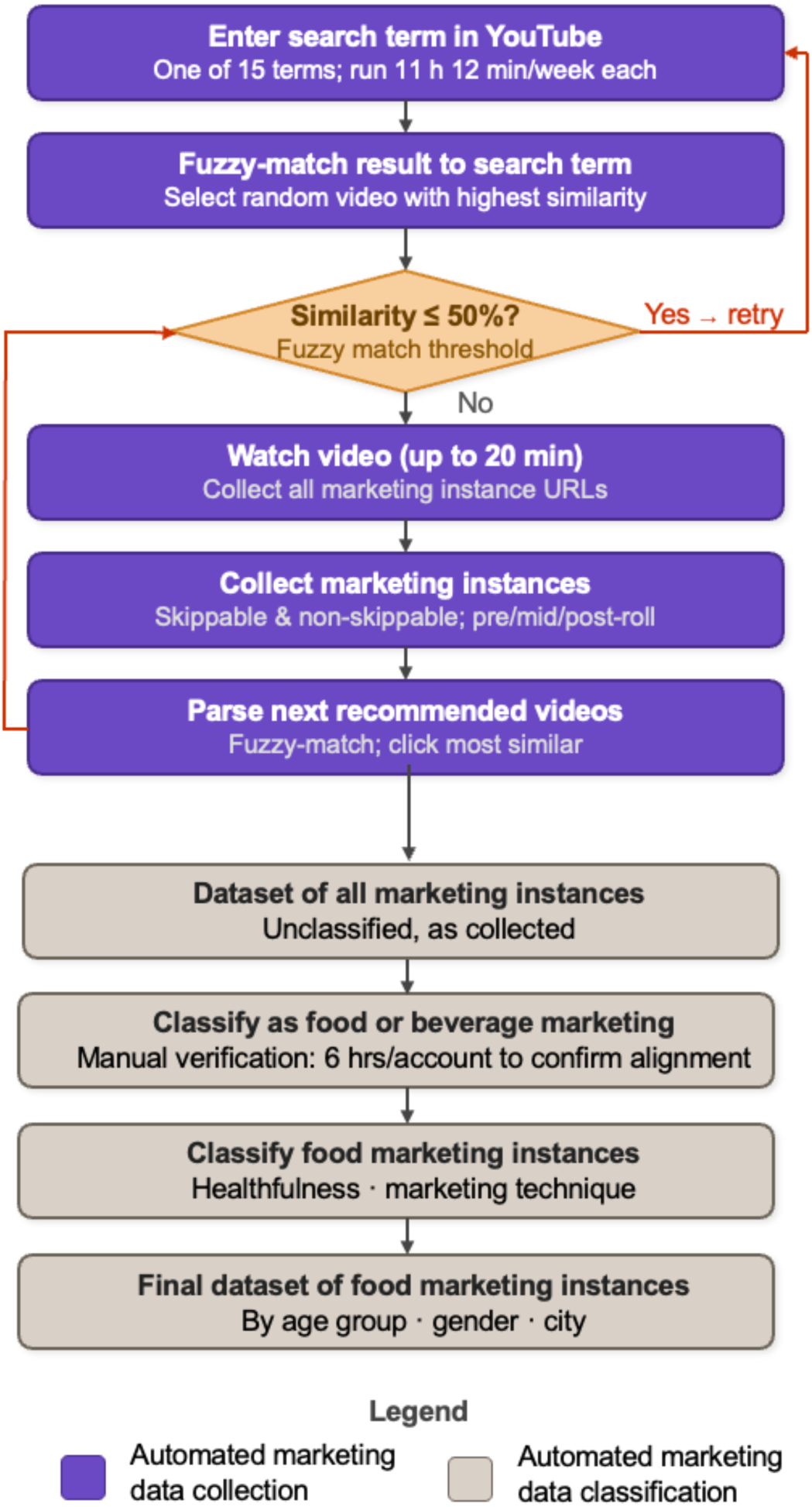
Overview of the CHOPPED automated data collection and classification pipeline for YouTube food marketing surveillance

**Table 1.** Characteristics of Avatar accounts^a^.

| Account ID | Age Group | Gender<sup>b</sup> |
|---|---|---|
| 1 | 2-4 years | Girl |
| 2 | 2-4 years | Boy |
| 3 | 5-8 years | Girl |
| 4 | 5-8 years | Boy |
| 5 | 9-12 years | Girl |
| 6 | 9-12 years | Boy |
| 7 | ≥ 18 years | Woman |
| 8 | ≥ 18 years | Man |

<sup>a</sup> One Avatar account per age and gender group.
<sup>a</sup> Video data were collected equally across four cities, with 336 hours collected per city for each account (1344 total hours per account).

For each account, 15 search terms were employed that were tailored to the preferences of each of the eight accounts based on age and gender (e.g., Fortnite videos for the boy’s account aged 9-12 years, craft ideas for the girl’s account aged 9-12 years; Supplementary Tables 1-9, Additional File 1). These search terms were selected based on seven studies examining YouTube video preferences and digital media use among boys, girls, men and women across particular age groups between 2018 and 2022 [20–26]. These search terms were essential because the accounts initially lacked browsing history that the YouTube algorithm could use to suggest videos. By using a designated search term as an initial filter, this process continuously generated new browsing data that aligned with each account’s intended age and gender profile, enabling the YouTube algorithm to deliver appropriately tailored content.

CHOPPED collected all skippable (allows viewers to skip after 5 seconds) and non-skippable (must be watched before a video can be viewed) video marketing instances played before, during, and after each video and moved on to the next video recommended by YouTube’s algorithm for each of the eight accounts. All marketing instances were collected from Avatar accounts that were not signed into YouTube. Each account was assigned a dedicated device to prevent viewing and search histories from carrying over between accounts. Each account was permitted to accumulate a short-term browsing history as it searched and viewed videos, consistent with how YouTube’s algorithm adapts recommendations and marketing instance delivery in real time for users who are not signed in. This approach was necessary because during a pilot test, we found that no food marketing instances were delivered to signed-in child accounts due to YouTube’s policy which automatically redirects accounts for children under 13 years to YouTube Kids unless a parent explicitly enables a supervised experience on regular YouTube, an opt-in step our pilot accounts did not use [27].

The AI system began by searching a given term on each account’s search bar using YouTube’s built in search functionality. It then compared the video title to the search term and proceeded to select a recommended video from the list of results that was most similar to the search term using fuzzy matching, a technique used to identify strings or terms that are approximately similar rather than exact matches. After selecting a random recommended video with the highest similarity, the system viewed it for up to 20 minutes, or in full if the video was shorter, and collected the URL for all marketing instances in the video within the 20 minutes period, excluding marketing instances that appeared after the 20-minute cut off. A 20-minute limit was imposed to prevent long videos from dominating the viewing time for each search term. This approach assumed that children view each consecutive video without an ad-blocker. After the video was complete, it parsed through the titles of subsequent videos recommended by the algorithm, compared them to the search terms, and clicked consecutive videos that were most similar to the search terms. If the AI system encountered a video with a similarity percentage of ≤50%, it restarted the search from the search bar and selected a more relevant video. We confirmed that this limited browsing patterns that did not align with the intended preferences of each account by manually viewing 6 hours’ worth of videos shown to each of the eight accounts. Without these search terms, the algorithm tended to drift toward unrelated video recommendations which did not reflect the viewing patterns of each age and gender group.

The AI system ran a total of 15 unique search terms per week for each of the eight accounts, with each running for 11 hours and 12 minutes weekly, totalling 168 hours of continuous monitoring per week per account. This duration was chosen to ensure equal time allocation for each search term within the system’s 24-hour daily runtime (i.e., 168 hours/week ÷ 15 search terms). We collected marketing instances from 4 major cities across Canada using our Virtual Private Network, including Vancouver, Calgary, Toronto, and Halifax, allocating 7 consecutive days in February and 7 consecutive days in March for each city while repeating the same 15 weekly search terms for each city. For each account, the AI system collected 1344 hours of video data across four Canadian cities between February and March 2024. Data collection occurred equally across all four cities, with 336 hours recorded per city (24 hours/day for 7 consecutive days in each month). Across all eight accounts, the AI system collected 10,752 hours of video data (1,344 hours/account × 8 accounts = 10,752 hours) between February and March 2024. Data collection proceeded without interruption throughout the study period. Monthly estimates elsewhere in this study are based on a standardized 28-day month, consistent with the 4-week (2 weeks/city × 4 cities) data collection structure described above.

### Classification of Food Healthfulness, Marketing Techniques, and Child-Targeted Food Marketing Instances

A marketing instance was defined as a video advertisement of a product, service, or brand displayed before, during, or after a video. Food-related marketing instances were defined as those that promoted a food product, main dish, multiple food products, or a multi-component meal through images, text, or auditory cues [28,29]. For marketing instances promoting multiple food products or a multi-component meal, each individual product was evaluated separately. The system classified the healthfulness of foods using the University of Toronto Food Classification List that includes nutrition information for over 80,000 pre-packaged products and 20,000 restaurant menu items up to 2020 [30]. Foods were deemed ‘unhealthy’ if they contained added saturated fats, sodium, or free sugars that exceeded Health Canada’s thresholds for foods that would be restricted under their proposed policy to restrict unhealthy food marketing to children (Table 2) [28, 29]. Food-related marketing instances were classified as unhealthy if at least one of the promoted products was unhealthy. Only when all individual products were classified as healthy was the entire marketing instance considered healthy. Brand marketing instances where no food products were featured were excluded.

**Table 2.**
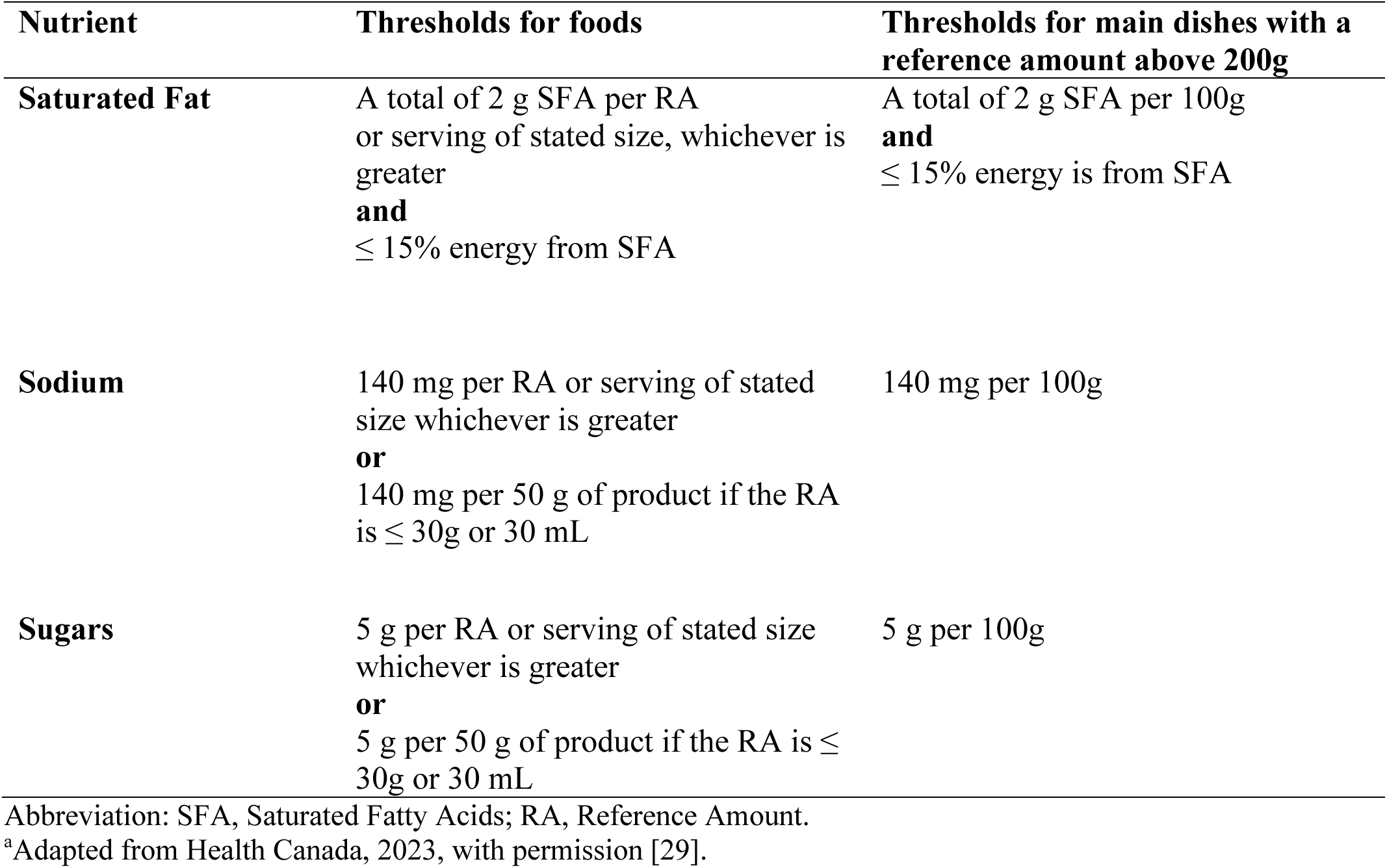
Health Canada’s proposed nutrient thresholds for restricting food marketing to children^a^.

For each marketing instance, the AI system also classified all of the marketing techniques present using Health Canada’s criteria for classifying food marketing techniques on digital media [28,29]. Health Canada’s marketing techniques include: presence of branded characters; presence of licensed characters; presence of celebrities/public figures; presence of other cartoon characters; child themes/visual design; use of child language; appeals to fun/cool; games and activities; price promotions, incentives, or giveaways; cross-promotions; unusual product flavour/shape; presence of children; adult-child situations; appeals to health/nutrition; displays of corporate and social responsibility; promotion of product convenience; appeals to flavour/texture; viral marketing; calls-to-action; special effects; and songs/music.

A child-targeted marketing instance was defined as a marketing instance that employed marketing techniques designed to capture the attention of children to influence their food preferences, desires, or purchasing intentions [28,29]. To identify child-targeted techniques, we considered a subset of Health Canada’s techniques based on the Child Appealing Packaging Tool [31] and a recent review that examined marketing techniques used to target children across multiple platforms [32]. Based on these studies, marketing techniques were classified as core child-targeted, broad child-targeted, or non-specific techniques as described in Supplementary Table 10, Additional File 1. We created these three classifications because Health Canada’s marketing techniques include techniques that may be present in the majority of marketing instances and were not categorized as child-targeted in previous research [28,29]. In the current study, marketing instances were considered child-targeted if they employed at least one core-child targeted marketing technique including: child themes/visual design; use of child language; appeals to fun/cool; games and activities; unusual product flavour, unusual product appearance; and adult-child situations. The presence of branded characters, licensed characters, celebrities/public figures, or other cartoon characters were excluded from the definition of core-child targeted techniques as CHOPPED did not have the capacity to capture these techniques at the time of the analyses. However, following the development of this aspect of the system, we subsequently conducted additional analyses that included these techniques, as reported in the sensitivity analyses.

The early version of CHOPPED used in this study was validated separately using a manually labelled sample of 647 marketing instances collected using the same child and adult accounts and search protocol described above. This validation sample was collected separately from the study dataset with an oversample of food marketing instances to ensure reliable estimation of sensitivity and specificity. Manual labels were assigned independently by two trained coders following a standardized coding manual, with disagreements resolved by consensus discussion. Briefly, CHOPPED demonstrated high sensitivity (0.84, 95% CI: 0.77–0.89) and specificity (0.83, 95% CI: 0.79–0.86) for identifying food marketing instances, and high sensitivity (0.85, 95% CI: 0.81–0.89) but lower specificity (0.58, 95% CI: 0.47–0.70) for identifying unhealthy food marketing instances. For the majority of core child-targeted marketing techniques (e.g., child themes/visual design, appeals to fun/cool, adult-child situations), the system showed high sensitivity but low specificity, indicating a tendency toward false positives rather than false negatives. Full performance metrics for this early version of the system are reported in Supplementary Table 11, Additional File 1.

### Data Analysis

To characterize the extent of unhealthy food marketing that targeted children, the total monthly number of unhealthy food marketing instances that used one or more core child-targeted techniques was expressed as frequencies and proportions of the total number of monthly food marketing instances collected from each of the eight accounts. To characterize the nature of unhealthy food marketing that targeted children, the total monthly number of unhealthy food marketing instances that used each core child-targeted technique was expressed as frequencies and proportions of the total number of monthly unhealthy food marketing instances that used any of the core-child targeted techniques collected from each of the eight accounts. Following further AI system development, the nature of unhealthy food marketing that targeted children was also characterized based on the presence of branded characters, licensed characters, celebrities/public figures, or other cartoon characters in a subsample of the data.

Descriptive statistics including frequencies and proportions of the extent and nature of food marketing were calculated. Poisson regression models were performed to assess whether the mean monthly frequencies of: food marketing instances, unhealthy food marketing instances, healthy food marketing instances, child-targeted food marketing instances, child-targeted unhealthy food marketing instances, and the use of each of the marketing techniques differed across the eight accounts. The unit of observation was the monthly frequency count for each account in each of the 2 months of data collection (February and March), yielding two observations per account (n=16 total). The two accounts representing children aged 2–4 years, which recorded zero food marketing instances in both months, were retained in all models as valid zero-count observations. Given the potential for overdispersion in marketing exposure count data, negative binomial regression was used in place of Poisson regression for all models; standard errors were clustered by account to account for the two repeated monthly observations contributed by each account. For each classification, we modelled the mean monthly frequency count (Yi) as a function of the intercept (β0), age (β1), gender (β2), and the interaction between age and gender (β3). The general model for each classification was:

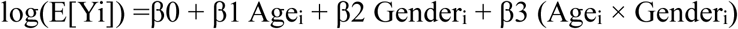

Pairwise comparisons of estimated marginal means were conducted following each Poisson regression model, with adjustments for multiple comparisons using Bonferroni corrections. Additionally, we calculated monthly means of the frequencies and proportions for both the extent and nature of food marketing across all child accounts combined and across all adult accounts combined. Monthly means were also presented based on a 2-hour daily viewing period by dividing the model-predicted monthly means by 12 periods (number of 2-hour viewing periods in a 24-hour day) to reflect the average time children typically spend on YouTube per day [10]. As actual 2-hour viewing periods were not directly observed, these values represent estimated 2-hour averages derived from monthly model estimates, rather than observed 2-hour exposure. T-tests were conducted to examine differences in mean monthly frequencies between child and adult accounts. All analyses were performed in R (version 4.3.0) [33], with p<0.05 indicating statistically significant differences.

### Sensitivity analyses

Sensitivity analyses were conducted to assess the impact of including the marketing techniques outlined by Health Canada that we did not classify as core child-targeted. First, the frequency and proportion of all unhealthy marketing instances that were child-targeted were calculated and compared to those from our primary analysis under two alternative definitions of child-targeted food marketing: using core and broad child-targeted techniques; and using core and broad child-targeted techniques along with non-specific techniques. Because the same marketing instances were reclassified under each of the three nested definitions rather than representing independent samples, Friedman tests were used to compare mean monthly frequencies of child-targeted unhealthy food marketing instances across the three definitions, with pairwise Wilcoxon signed-rank tests used for post-hoc comparisons where the omnibus test was significant.

In the second sensitivity analysis, we calculated the frequency of child-targeted food marketing instances and then applied the same approach to examine the impact on our findings when branded characters, licensed characters, celebrities/public figures, and other cartoon characters were included as core-child targeted techniques and compared to the two alternative definitions of child-targeted food marketing: using core and broad child-targeted techniques; and using core and broad child-targeted techniques along with non-specific techniques. These frequencies were also compared within each definition when including and excluding the presence of characters and celebrities/public figures. This analysis was performed on a smaller subset of the available data (n= 2,599 food marketing instances) because the corresponding videos of some marketing instances were no longer available on YouTube at the time of the analysis.

## RESULTS

### The Extent of Food Marketing Instances and Unhealthy Food Marketing Instances

Over 2 months, CHOPPED collected 60,633 marketing instances from all eight accounts, of which 11,324 were unique. Of the total marketing instances, 8.7% (n=5,263) were food-related. There were significant differences in the monthly frequency of food marketing instances between some accounts when considering a 24-hour daily viewing period (Table 3). Over a 24-hour daily viewing period, the monthly frequency (%) of food marketing instances shown to the boy’s account aged 9-12 years (485, 9.5%) was significantly higher than that shown to the boy’s account aged 5-8 years (364, 8.3%) and the man’s account (388, 8.1%). The monthly frequency (%) of food marketing instances shown to the boy’s account aged 5-8 years (364, 8.3%) was significantly lower than that shown to the girl’s account aged 9-12 years (486, 8.9%) and the woman’s account (480, 9.7%). The monthly frequency of food marketing instances shown to the man’s account (388, 8.1%) was significantly lower than that shown to the girl’s account aged 9-12 years (486, 8.9%) and the woman’s account (480, 9.7%). However, these monthly differences were not significant when considering a 2-hour daily viewing period. The monthly frequency of food marketing instances was similar for all other accounts for both viewing periods. Additionally, no food marketing instances were shown to child accounts between the ages of 2-4 years.

**Table 3.** The extent of food marketing shown to child and adult accounts by age and gender.

| Classification | Mean monthly frequency (%) based on a 24-hour or a 2-hour daily viewing period |  |  |  |  |  |  |  |  |  |  |  |
|---|---|---|---|---|---|---|---|---|---|---|---|---|
|  | Boy<br>5-8 years |  | Girl<br>5-8 years |  | Boy<br>9-12 years |  | Girl<br>9-12 years |  | Man<br>≥ 18 years |  | Woman<br>≥ 18 years |  |
|  | 24-hours | 2-hours | 24-hours | 2-hours | 24-hours | 2-hours | 24-hours | 2-hours | 24-hours | 2-hours | 24-hours | 2-hours |
| **Food marketing instances** | 364<sup>a</sup><br>(8.3) | 30<br>(8.3) | 430<br>(7.6) | 36<br>(7.6) | 485<br>(9.5) | 40<br>(9.5) | 486<sup>b, c</sup><br>(8.9) | 41<br>(8.9) | 388<sup>a</sup><br>(8.1) | 32<br>(8.1) | 480<sup>b, c</sup><br>(9.7) | 40<br>(9.7) |
| **Unhealthy food marketing instances** | 253<sup>a</sup><br>(69.5) | 21<br>(70.0) | 281<sup>a</sup><br>(65.4) | 23<br>(63.9) | 367<br>(75.7) | 31<br>(77.5) | 302<br>(62.4) | 25<br>(61.0) | 242<sup>a</sup><br>(62.4) | 20<br>(62.5) | 292<sup>a</sup><br>(60.9) | 24<br>(60.0) |
| **Healthy food marketing instances** | 111<br>(30.5) | 9<br>(30.0) | 149<br>(34.6) | 12<br>(33.3) | 118<br>(24.3) | 10<br>(25.0) | 182<sup>a, b</sup><br>(37.6) | 15<br>(36.6) | 146<br>(37.6) | 12<br>(37.5) | 188<sup>a, b</sup><br>(39.1) | 16<br>(40.0) |
| **Child-targeted food marketing instances** | 347<sup>a</sup><br>(95.3) | 29<br>(96.7) | 400<br>(93.0) | 33<br>(91.7) | 456<br>(94.0) | 38<br>(95.0) | 464<sup>b, c</sup><br>(95.0) | 39<br>(95.1) | 365<sup>a</sup><br>(94.1) | 30<br>(93.8) | 443<sup>b</sup><br>(92.3) | 37<br>(92.5) |
| **Child-targeted unhealthy food marketing instances** | 241<sup>a</sup><br>(66.2) | 20<br>(66.7) | 261<sup>a</sup><br>(60.6) | 22<br>(61.1) | 340<br>(70.2) | 28<br>(70.0) | 289<br>(59.5) | 24<br>(58.5) | 224<sup>a</sup><br>(57.7) | 19<br>(59.4) | 263<sup>a</sup><br>(54.7) | 22<br>(55.0) |

Analyses were conducted using negative binomial regression followed by pairwise comparisons of estimated marginal means to examine differences in frequency by age and gender.
The denominator for calculating the proportion of food marketing instances is the total monthly frequency of both food and non-food marketing instances shown to each account. The denominator for calculating the proportion of all other classifications is the total monthly frequency of food marketing instances shown to each account.
The total monthly viewing time per account was 672 hours (24 hours x 7 days x 4 weeks) for a continuous 24-hour daily viewing period and 56 hours (2 hours x 7 days x 4 weeks) for a 2-hour daily viewing period.
The total monthly frequency of food marketing instances collected from all eight accounts was 2,632, while the total monthly frequency of non-food marketing instances was 27,685.
No food marketing instances were shown to the child accounts 2-4 years.
<sup>a</sup> Significantly different than the boy 9-12 years account (p < 0.05).
<sup>b</sup> Significantly different than the boy 5-8 years account (p < 0.05).
<sup>c</sup> Significantly different than the man’s account (p < 0.05).

The mean monthly frequency (%) of food marketing instances shown to all child accounts combined did not differ significantly from those shown to all adult accounts during the 24-hour (Child: 441, 8.6%; Adult: 434, 8.9%) or the 2-hour (Child: 37, 8.6%; Adult: 36, 8.9%) daily viewing periods (Figure. 2).

**Figure 2.**
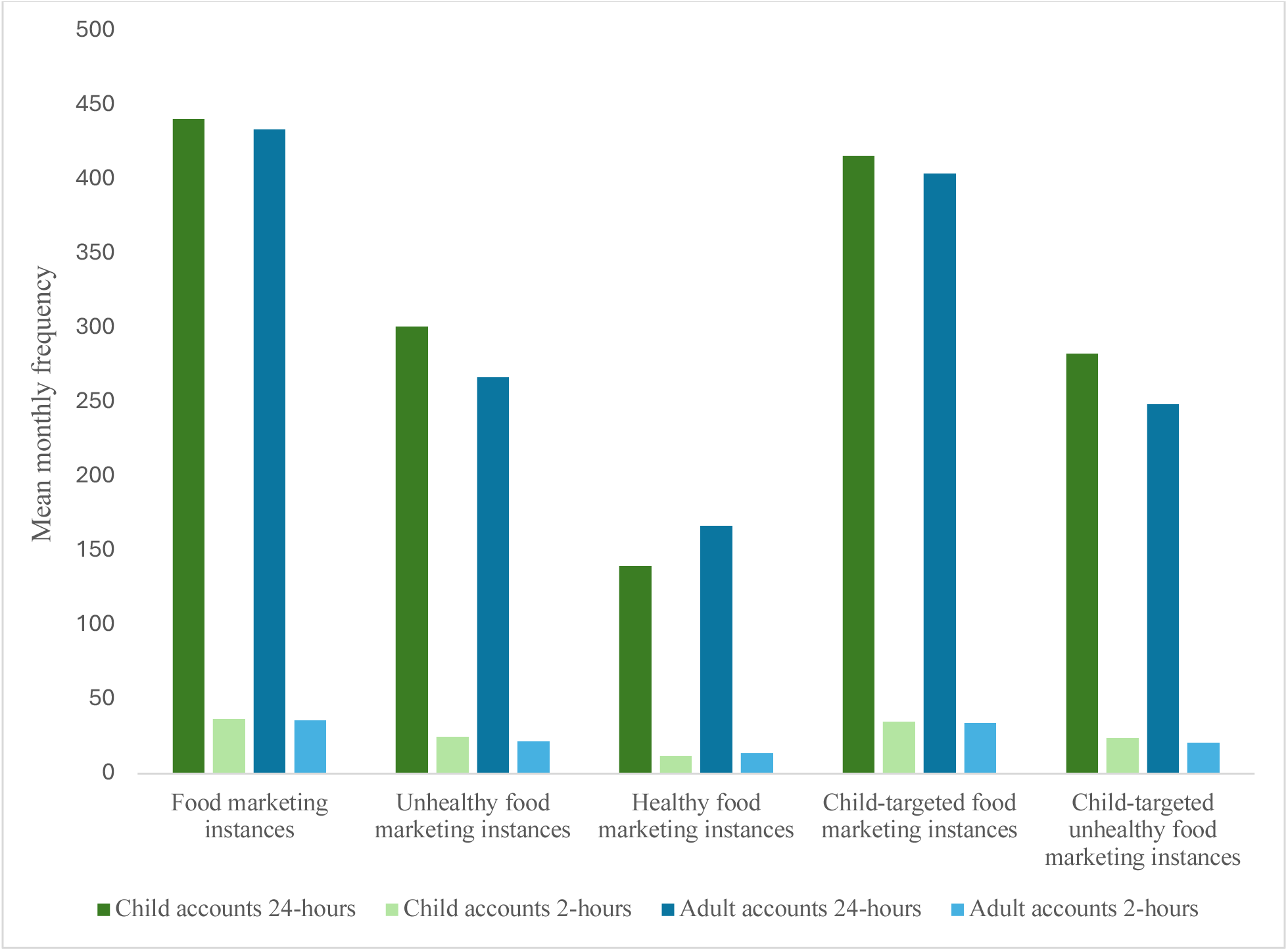
The extent of food marketing instances shown to all child and adult accounts.^a,b^ Analyses were conducted using t-tests to examine differences in mean monthly frequencies between child and adult accounts. No significant differences were found between child and adult accounts during a 24-hour or a 2-hour daily viewing period. ^a^Mean monthly frequency for the four child and two adult accounts that were shown food marketing. ^b^No food marketing instances were shown to the child accounts 2-4 years.

There were also significant differences in the monthly frequency of unhealthy food marketing instances between some accounts (Table 3). The monthly frequency (%) of unhealthy food marketing instances shown to the boy’s account aged 9-12 years (367, 75.7%) was significantly higher than that shown to the boy’s account aged 5-8 years (253, 69.5%), the girl’s account aged 5-8 years (281, 65.4%), the man’s account (242, 62.4%), and the woman’s account (292, 60.9%), (Table 3). However, these monthly differences were not significant when considering a 2-hour daily viewing period. The monthly frequency of unhealthy food marketing instances was similar for all other accounts for both viewing periods.

The mean monthly frequency (%) of unhealthy food marketing instances shown to all child accounts combined did not differ significantly from those shown to all adult accounts during the 24-hour (Child: 301, 68.3%; Adult: 267, 61.5%) or the 2-hour (Child: 25, 67.6%; Adult: 22, 61.1%) daily viewing periods (Figure. 2).

### The Extent of Child-Targeted Unhealthy Food Marketing Instances

Over a 24-hour daily viewing period, the mean monthly frequency (%) of child-targeted unhealthy food marketing instances shown to the boy’s account aged 9-12 years (340, 70.2%) was significantly higher than that shown to the boy’s account aged 5-8 years (241, 66.2%), girl’s account aged 5-8 years (261, 60.6%), the man’s account (224, 57.7%), and the woman’s account (263, 54.7%) (Table 3). These monthly differences were not significant when considering a 2-hour daily viewing period. The monthly frequency of child-targeted unhealthy food marketing instances was similar for all other accounts for both viewing periods.

The mean monthly frequency (%) of child-targeted unhealthy food marketing instances shown to all child accounts combined did not differ significantly from those shown to all adult accounts during the 24-hour (Child: 283, 64.2%; Adult: 244, 57.4%) or the 2-hour (Child: 24, 64.2%; Adult: 21, 57.4%) daily viewing periods (Figure. 2).

### The Nature of Child-Targeted Unhealthy Food Marketing Instances Using Core Child-Targeted Techniques

Multiple core child-targeted techniques were used in unhealthy food marketing instances (Table 4 and Figure. 3). Some core child-targeted techniques were shown at significantly different frequencies across accounts. For instance, the boy’s account (9-12 years) was shown a higher monthly frequency of adult-child situations, child language, child themes/visual design, appeals to fun/cool compared to the boy’s (5-8 years), girl’s (5-8 years), woman’s, and man’s accounts (Table 4). For the technique appeals to fun/cool, the boy’s account (9-12 years) was shown a higher monthly frequency compared to the girl’s account (9-12 years). The man’s account was shown a lower monthly frequency of the techniques child themes/ visual design and unusual product appearance than the girl’s account (9-12 years). Similarly, the man’s account was shown a lower monthly frequency of the technique unusual product appearance compared to the woman’s account. However, these differences were not significant during a 2-hour daily viewing period. The most frequently used core-child targeted marketing techniques for child and adult accounts were similar and included adult-child situations, use of child language, child themes/visual design, unusual product appearance, and appeals to fun/cool. By contrast, unusual product flavour and games and activities were rarely shown to child or adult accounts and did not significantly differ across accounts.

**Figure 3.**
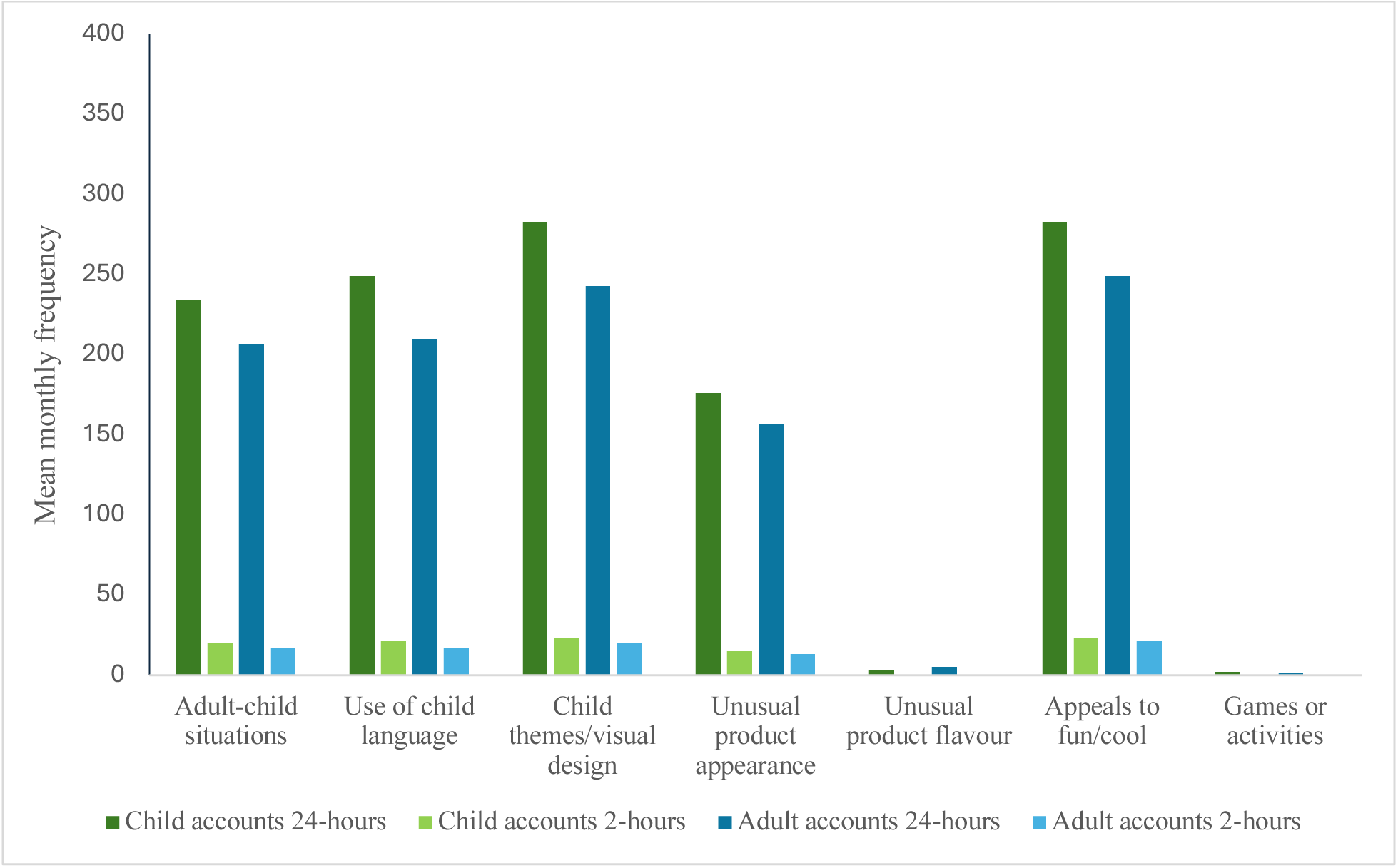
The nature of child-targeted unhealthy food marketing instances (using core child-targeted techniques) shown to all child and adult accounts.^a,b^ Analyses were conducted using t-tests to examine differences in mean frequencies between child and adult accounts. No significant differences were observed between child and adult accounts in the mean monthly frequency of unhealthy food marketing instances that used any of the core child-targeted marketing techniques during a 24-hour daily viewing period or a 2-hour daily viewing period. ^a^ Mean monthly frequency for the four child and two adult accounts that were shown food marketing. ^b^ No food marketing instances were shown to the child accounts 2-4 years.

**Table 4.**
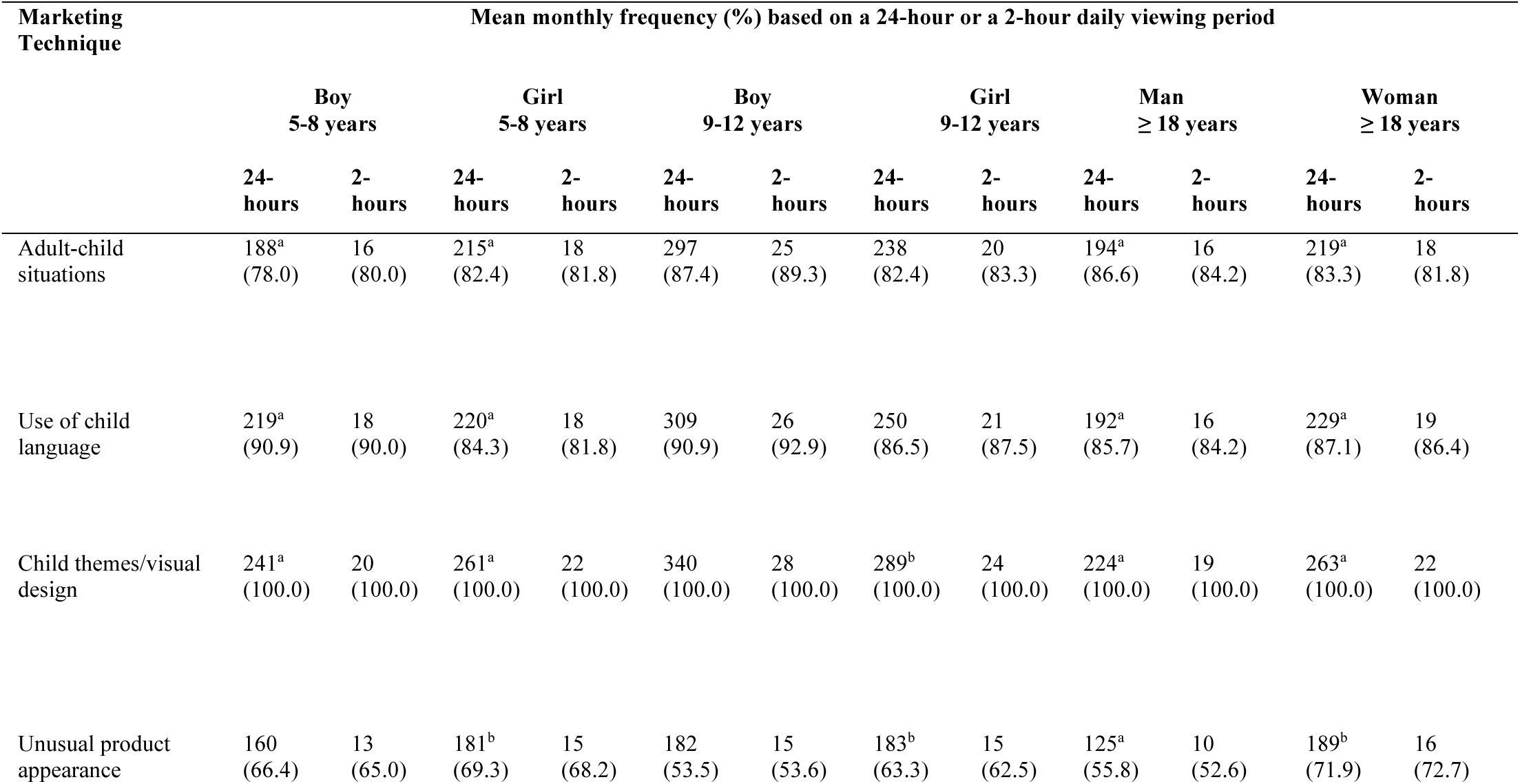
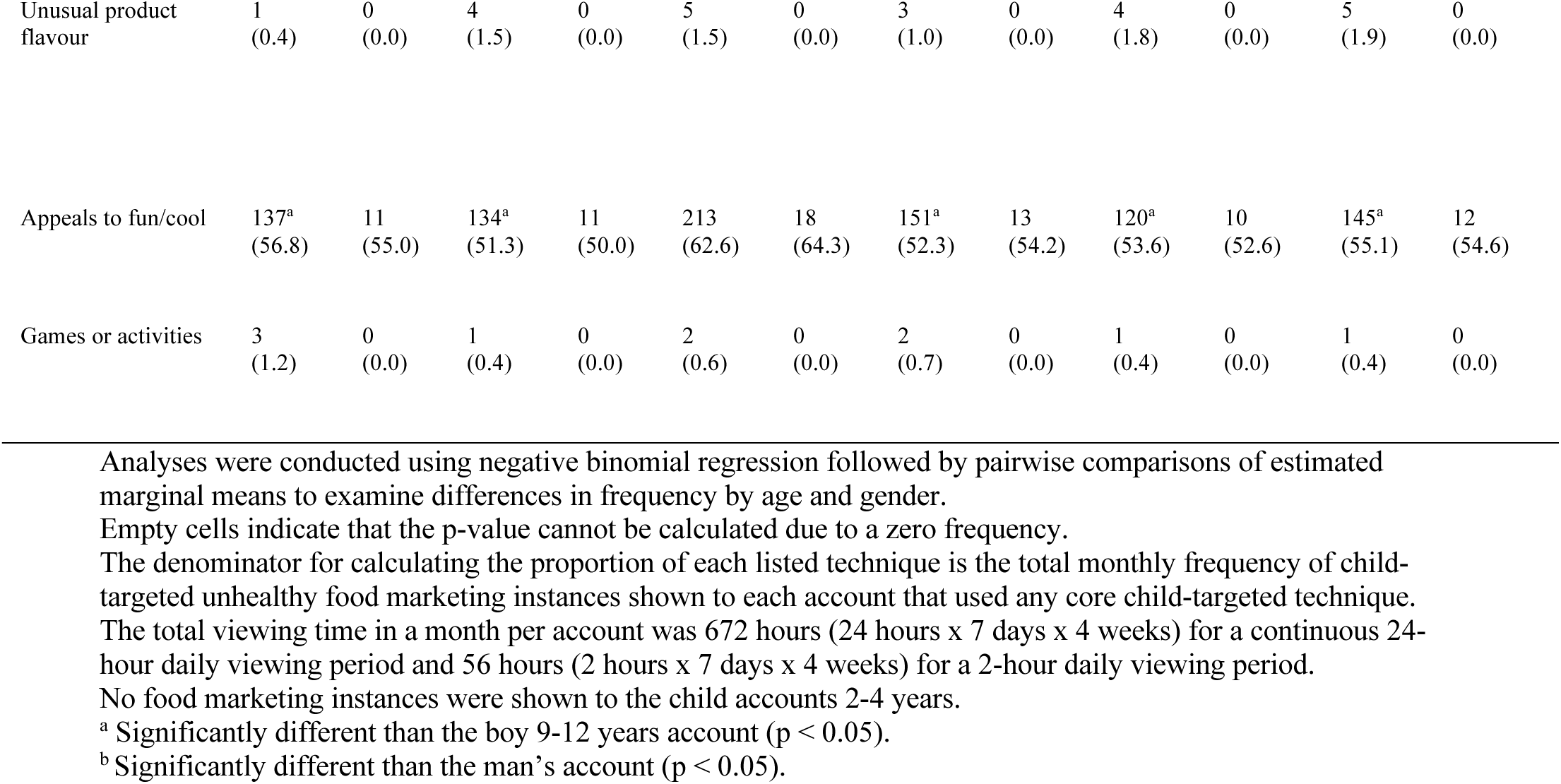
The nature of child-targeted unhealthy food marketing instances (using core child-targeted techniques) shown to child and adult accounts by age and gender.

### Sensitivity Analyses

In sensitivity analyses, we compared two alternative definitions of child-targeted unhealthy food marketing to our primary definition which included only core child-targeted techniques (Supplementary Figure 1 and 2, Additional File 1). Under our primary definition, almost all unhealthy food marketing instances were considered child-targeted (child accounts: 283, 94.0%; adult accounts: 243, 91.0%). Under the first alternative definition which included core and broad child-targeted techniques, we found that the mean monthly frequency and proportion of child-targeted unhealthy food marketing were identical to results from our primary analyses (child accounts: 283, 94.0%; adult accounts: 243, 91.0%; p > 0.05). However, under the second definition which included core, broad, and non-specific techniques, all unhealthy food marketing instances were classified as child-targeted (child accounts: 301, 100.0%; adult accounts: 267, 100.0%; p > 0.05). These frequencies did not significantly differ from those in our primary analyses or when using the first alternative definition.

In the second sensitivity analysis of a subsample of 2,599 food marketing instances (23.0% of total sample), the presence of branded characters, licensed characters, celebrities/public figures, or other cartoon characters was very low across the subsample with proportions ranging from 1.0% to 16.0% during a 24-hour viewing period, with branded characters being the most prevalent and licensed characters the least (Supplementary Table 12, Additional File 1). The prevalence of these characters was not significantly different across accounts.

When the presence of characters and celebrities/public figures were included as child-targeted techniques, the majority of unhealthy food marketing instances in the subsample were considered child-targeted under our primary definition (child accounts: 145, 79.2%; adult accounts: 151, 77.4%) and under the first alternative definition (child accounts: 145, 79.2%; adult accounts: 151, 77.4%) with no differences between them (Supplementary Figure 2, Additional File 1). Under the second alternative definition, all unhealthy food marketing instances were classified as child-targeted (child accounts: 183, 100.0%; adult accounts: 195, 100.0%). These frequencies did not significantly differ from those in our primary analyses or when using the first alternative definition. Additionally, there were no significant differences in the frequency of child-targeted marketing instances within each definition when including or excluding the presence of characters and celebrities/public figures (Supplementary Figure 2, Additional File 1 ). Therefore, the classification of unhealthy food marketing as child-targeted was not meaningfully influenced by whether the presence of characters or celebrities/public figures was included as a qualifying criterion, supporting the robustness of our main findings.

## DISCUSSION

This study pioneered the use of AI to assess the extent and nature of unhealthy food marketing that children may encounter on YouTube. Of all the food marketing instances shown to child accounts, 64% were child-targeted marketing instances for unhealthy foods. Five key findings emerged from this research. First, child accounts between 5-12 years were shown a mean of 25 unhealthy food marketing instances per month, or less than one per day during children’s typical 2-hour daily viewing period on YouTube. Second, younger child accounts (aged 2-4 years) were not shown any food marketing instances. Third, the boy’s account 9-12 years was shown more monthly instances of child-targeted unhealthy food marketing and of some core child-targeted techniques than most other accounts based on a 24-hour time frame. Fourth, the inclusion of characters and celebrities/public figures did not change our findings. Lastly, most or all unhealthy food marketing instances were classified as child-targeted regardless of the definition used.

Our first key finding was that child accounts between 5-12 years were shown 25 unhealthy food marketing instances per month, or less than one per day during a 2-hour daily viewing period. Twenty four of these marketing instances were child-targeted. Our AI-based analysis showed lower monthly potential exposure to unhealthy food marketing compared to previous studies. One prior study that used manual methods to examine children’s exposure to unhealthy food marketing on YouTube estimated that children may be exposed to 44 unhealthy food marketing instances per month for the same viewing period of 2-hours per day [13]. Our results suggest a monthly potential exposure that is somewhat lower than the previously reported estimate.

Moreover, given that the AI system had a low specificity in detecting unhealthy food marketing instances and a low sensitivity in identifying healthy food marketing instances we may have overestimated children’s potential exposure. This inconsistency in estimates of the frequency of unhealthy food marketing instances may be explained by differences in methodology. First, the prior study manually monitored potential exposure using incognito mode, which does not account for personalized algorithms and likely does not capture the tailored marketing content that children may encounter, possibly misestimating children’s potential exposure. Additionally, the prior study was conducted over a limited period of 3 weeks, with only 54 hours and 35 minutes of recorded video, offering a limited view of the marketing landscape. By comparison, we collected 5,376 hours of marketing data over 2 months across the four child accounts that were shown ads. It is also important to note that our study did not specify the exact timing of the 2-hour viewing period. Instead, we assumed that the frequency of marketing instances remained consistent throughout a 24-hour period and proportionally calculated the mean number of marketing instances for a 2-hour window. Previous studies have shown that marketing may fluctuate based on the time of day [34], potentially driving this inconsistency in findings.

However, it is possible that the difference in estimates could be entirely due to the fact that the prior study was conducted in Malaysia, whereas our study was conducted in Canada. Previous studies have shown that children’s food marketing exposure varies by country [35]. Nevertheless, while we found that children’s exposure to unhealthy food marketing on YouTube may be relatively low, YouTube represents only one digital media platform on which children may encounter such marketing and exposure across platforms is cumulative. Moreover, a high proportion of the instances observed were child-targeted, which could potentially have a negative impact on children’s dietary patterns [12].

Our second key finding was that younger child accounts (aged 2-4 years) were not shown any food marketing instances. This is likely because the search terms that were used for these accounts allowed YouTube’s algorithm to identify these accounts as belonging to young children. As a result, the algorithm redirected these accounts to videos with YouTube’s “made-for-kids” designation, which prohibits food marketing on these videos [27]. Our findings do not align with previous research that found food marketing instances displayed before, during, and after popular “made-for-kids” videos on YouTube [37]. However, that study was conducted before YouTube implemented its new regulations which restrict food marketing on “made-for-kids” content [27]. Thus, our findings suggest that YouTube’s current protective measure, the "made-for-kids" designation, may effectively protect younger children (aged 2-4 years) from food marketing, but leaves older children (aged 5-12 years) exposed to unhealthy food marketing. Importantly, this finding should be interpreted with caution, as in real-world settings young children may be exposed to marketing through shared devices or by using others’ accounts. If researchers and policymakers only monitor unhealthy food marketing on “made-for-kids” videos, they risk underestimating children’s potential exposure to unhealthy food marketing, as older children often watch general-audience content [23,24].

Our third key finding was that the boy’s account (9-12 years) was shown more monthly unhealthy food marketing instances and more child-targeted unhealthy food marketing instances than most other accounts based on a 24-hour time frame. Moreover, these marketing instances more often included adult-child situations, child language, child themes/visual design, and appeals to fun/cool. However, these differences were not significant when considering children’s typical viewing period of 2-hours per day and may therefore lack public health relevance as it is unrealistic for a child to spend 24 hours a day watching YouTube. Additionally, the AI system’s low specificity in identifying unhealthy food marketing instances suggests that the frequency of monthly unhealthy food marketing instances may be overestimated. Our findings do not align with a previous study in the UK, which found that a child account 9-12 years was shown a similar number of child-targeted unhealthy food marketing as an adult account on YouTube over a 24-hour monitoring period [36]. This discrepancy may be explained by differences in the monitoring period which was limited to one week in the UK study or by variations in how marketers target children across the UK and Canada. With respect to our findings that the boy’s account was shown marketing instances that used different marketing techniques compared to other accounts, a previous study showed that exposure to child-targeted marketing techniques on television was higher for girls than for boys in Calgary, Canada, especially for the marketing technique of use of child language [38]. Another study on social media that included children between 12-16 years found that there was no difference in the frequency of food marketing instances shown to boys and girls [39]. These differences in findings could be due to differences in age and gender targeting on different platforms. Overall, although we found some differences in the frequency and nature of unhealthy food marketing shown to the older boy’s account, the extent and nature of unhealthy food marketing during typical use appears relatively similar by age and gender on YouTube. This could indicate that food marketing on YouTube aims to maximize overall reach and may not target children by age or gender.

Our fourth key finding was that the inclusion of characters and celebrities/public figures did not change our findings. Although characters and celebrities/public figures are widely cited as prominent child-targeted marketing techniques [32], they appeared in only 1.0 to 16.0% of child-targeted unhealthy food marketing instances in this study. Consistent with this, sensitivity analyses confirmed that including or excluding these techniques did not meaningfully change the frequency of child-targeted unhealthy food marketing instances identified, suggesting that excluding them from our primary analysis did not meaningfully affect our findings. Our findings are broadly consistent with a recent study that similarly found that character-based techniques accounted for approximately 8% of food marketing instances across multiple digital media platforms [14].

Our last key finding was that most or all unhealthy food marketing instances were classified as child-targeted regardless of the definition we used. The frequency of child-targeted unhealthy food marketing did not change whether we considered only core child-targeted techniques or included both core and broad techniques. This was because the inclusion of broad techniques did not introduce additional instances classified as child-targeted, resulting in an unchanged overall frequency of child-targeted food marketing instances. When non-specific techniques were included, all marketing instances were classified as child-targeted since the technique calls-to-action appeared in every food marketing instance. However, the AI system’s lower performance in identifying broad child-targeted marketing techniques and mixed performance for non-specific techniques may have rendered some aspects of our sensitivity analyses less meaningful.

This study has several limitations. First, although we used evidence-based search terms to simulate browsing patterns based on age and gender, these terms may not fully reflect the search habits of Canadian users. We also only captured video marketing instances and excluded image marketing, banner marketing, in-feed marketing, influencer marketing, and brand marketing instances, possibly underestimating children’s potential exposure to food marketing on YouTube. Additionally, we analyzed YouTube using non-signed-in accounts. If children were signed in, they would not be exposed to any food marketing instances. CHOPPED assessed the healthfulness of foods using the 2020 version of the University of Toronto Food Classification List [28], possibly leading to some misclassification of food healthfulness if some foods have since been reformulated. However, this is unlikely to have materially affected our findings as most unhealthy food marketing was for items that were clearly unhealthy and unlikely to have significantly changed in nutritional composition over time (e.g., soda, chocolate bars, confectionary). Data collection spanned 2 months and therefore cannot fully represent children’s potential longer-term exposures to food marketing on YouTube. However, we collected data for 10,752 hours for all eight child and adult accounts combined (i.e., 8,064 hours for child and 2,688 hours for adult accounts) over a period of 2 months which is substantially longer than prior studies that collected no more than 336 hours of food marketing data on YouTube [13,36].

Additionally, our AI system collected 60,633 total marketing instances, 11,324 of which were unique. These values far surpass those of previous studies which collected at most 187 unique marketing instances on YouTube [13,36]. Finally, based on performance metrics, CHOPPED may have overestimated children’s potential exposure to unhealthy child targeted food marketing instances. Although CHOPPED could not identify branded and licensed characters, celebrities/public figures, or other cartoon characters children at the time of this analysis, it does not appear that this limitation influenced our findings. The sensitivity analysis did not show a significant difference in the extent or nature of child-targeted marketing instances when including versus excluding these techniques in the definition of child-targeted marketing.

## CONCLUSION

In conclusion, our findings suggest that the frequency of unhealthy food marketing shown to children on YouTube may be low, with less than one unhealthy food marketing instance per day over a 2-hour daily viewing period. Although we found some evidence that the older boys’ account was shown more monthly instances of child-targeted unhealthy food marketing and of some core child-targeted techniques than other child and adult accounts, these differences may be of limited public health relevance. Overall, there was limited evidence that marketing targeted children by age or gender on YouTube. As the first study to use AI to monitor food marketing to children on digital media, we demonstrated that AI can effectively monitor the digital food marketing landscape over extended periods. Our findings highlight the potential of AI-driven surveillance to inform policies to restrict unhealthy food marketing to children, which may help to protect children from the negative impacts of unhealthy food marketing.

## Data Availability

All data produced in the present study are available upon reasonable request to the authors

## ABBREVIATIONS

AI: Artificial Intelligence
CHOPPED: CHild Oriented food Product Promotion Evaluation and Detection system
US: United States
UK: United Kingdom

## DECLARATIONS

### Ethics Approval and consent to participate

Not applicable.

### Consent to Participate

Not applicable.

### Consent for publication

Not applicable.

### Availability of data and materials

The datasets used and/or analyzed during the current study are available from the corresponding author on reasonable request.

### Author’s contributions

SAS, LRP, DLO, MPK, and CE conceptualized and designed the study. SAS, TW, SF, IL, and DLO designed and trained the AI system. SAS, SY, and SF collected and analyzed the data. All authors interpreted the data, critically edited the manuscript, approved the final version, and agreed to be accountable for all aspects of the work.

