## Supplementary Tables 1-9; Supplementary Table 10; Supplementary Table 11; Supplementary Figure 1 and 2; Supplementary Figure 2; Supplementary Table 12 for "Unboxing unhealthy food marketing: An analysis of the extent and nature of unhealthy food marketing to children on YouTube in Canada using the CHOPPED artificial intelligence system"

**Additional Files**

*Supplemental Tables*

**Table 1** Studies used to inform search terms and their description

| Study | Description |
|---|---|
| 1.Radesky, J. S., Schaller, A., Yeo, S. L., Weeks, H. M., & Robb, M.B. Young kids and YouTube: How ads, toys, and games dominate viewing. Common Sense Media. 2020. https://www.commonsensemedia.org/sites/default/files/research/report/2020_youngkidsyoutube-report_final-release_forweb_1.pdf. Accessed 25 Jan 2025. | This study examined YouTube use among children aged 0 to 8 years in the US between March 26 and April 1, 2020. A total of 191 parents provided a list of the last videos their children watched on the main YouTube site. |
| 2.Rideout, V., & Robb, M. B. The Common Sense census: Media use by kids age zero to eight, 2020. Common Sense Media. 2020 https://www.commonsensemedia.org/sites/default/files/research/report/2020_zero_to_eight_census_final_web.pdf. Accessed 25 Jan 2025. | This online survey was conducted with 1,440 parents of children aged 8 years or younger from February 18 through March 13, 2020, in the US. Parents reported the frequency of children's use of various types of media, the amount of time spent doing various media activities, and how media content was selected. |

| | |
|---|---|
| 3.Rideout, V., Peebles, A., Mann, S., & Robb, M. B. Common Sense census: Media use by tweens and teens. Common Sense Media. 2022. https://www.commonsensemedia.org/sites/default/files/research/report/8-18-census-integrated-report-final-web_0.pdf. Accessed 25 Jan 2025. | This online survey was conducted with 1,306 children and teens aged 8 to 18 years in 2019 and 2021 in the US. The survey asked children and teens questions about their preferred digital media activities and the amount of time spent on each digital media platform. |
| 4.Rollins, D., Bridgewater, E., Munzer, T., Weeks, H. M., Schaller, A., Yancich, M., Gipson, W., Drogos, K., Robb, M. B., & Radesky, J.S. Who is the "you" in YouTube? Missed opportunities in race and representation in children's YouTube videos. Common Sense Media . 2022. https://www.commonsensemedia.org/sites/default/files/research/report/2022-youtube-report-final-web.pdf. Accessed 25 Jan 2025. | This study examined YouTube use among 114 children (0 to 8 years) between March and April 2020, and 140 teens (9 to 18 years) between June and July 2021. Parents of children provided a list of the last videos their children had watched on the main YouTube site, while teens provided a list of the last videos they had watched on YouTube. |
| 5. Scott, F., Parry, B., Marsh, J., Lahmar, J., Nutbrown, B., Scholey, E., Baldi, P., Law, L., & Yamada-Rice, D. Addressing the "Whys" of UK Children's YouTube Use: A Purposes Approach. Social Media + Society. 2023: 9:4. | This study examined 3,154 children and adolescents' (aged from 0 to 16 years) use of social media (YouTube and YouTube kids were included as social media platforms) and television. Online surveys were conducted with 3,154 families with children across the UK. Six case studies were also conducted with families with 14 children aged 2 to 14 years (8 girls and 6 boys) to describe the role of television and digital media in each family's life. Quantitative and qualitative results highlighted YouTube videos that children in various age groups watched. |

| | |
|---|---|
| 6.Children and parents: media use and attitudes report. 2023. https://www.ofcom.org.uk/__data/assets/pdf_file/0027/255852/childrens-media-use-and-attitudes-report-2023.pdf. Accessed 25 Jan 2025. | This study examined digital media use among 6,795 children and adolescents between 3 and 17 years of age. Media included Facebook, Instagram, YouTube, Snapchat, TikTok, WhatsApp, television, and video games. Surveys were conducted with children and adolescents between May 2022 and November 2022 in the UK. |
| 7.Life on the small screen: What children are watching and why. 2019. https://www.ofcom.org.uk/__data/assets/pdf_file/0021/134832/Ofcom-childrens-content-review-Publish.pdf. Accessed 25 Jan 2025. | This study examined digital media use among 40 children and adolescents between 4 and 16 years of age. Media included YouTube, television, and streaming apps. This study was conducted between October and December 2018 in the UK. |
| 8.Landrum AR. Are Women a Missing Audience for Science on YouTube? An Exploratory Study. Front. Commun. 2021: Apr 6:610920. | This study conducted two surveys with 1,003 women and men aged between 18 and 85 years regarding their YouTube viewing habits. This study was conducted between 2019 and 2020 in the US. |

**Table 2** Search terms for boys between 9 and 12 years of age and evidence supporting each term

| Search Term | Evidence supporting the search term |
|---|---|
| Xbox games<br>PlayStation games<br>PC gaming<br>Fortnite videos<br>Parkour videos<br>Baseball videos<br>Basketball videos<br>Soccer videos<br>F2 videos<br>Football videos<br>Hockey videos<br>Tennis videos<br>WWE videos<br>Movie theories<br>Game theories | Boys between 9 and 12 years who participated in study #1 and #5 reported watching gaming-related videos, such as Xbox and PlayStation games, PC gaming, and Fortnite tutorials.[1,2] They also watched parkour videos, sports-related content like F2, movie theories, and game theories channels all on YouTube.[1,2] |

**Table 3** Search terms for girls between 9 and 12 years of age and evidence supporting each term

| Search term | Evidence supporting the search term |
|---|---|
| DIY videos<br>Funny videos<br>Music videos<br>90s music | Girls aged 9 to12 years who participated in study #5 said that they watched Do It Yourself (DIY) videos, humorous videos, music videos and channels (e.g., 90s music videos), lifestyle vloggers, autonomous sensory meridian response (ASMR) videos, science experiments, and cooking vlogs on YouTube.[2] |
| Piano tutorial<br>Guitar tutorial<br>Lifestyle vloggers<br>Cooking vlogs<br>Science experiments<br>ASMR videos | In study #7, girls (aged 9 to 12 years) reported that they watched guitar and piano tutorials on YouTube.[3] |
| Make-up tutorials<br>Craft ideas<br>Easy crafts<br>Musician interviews<br>Family videos | Girls between 9 and 12 years who participated in study #4 and #7 said that they watched make-up tutorials, crafts, musician interviews, and YouTube families on YouTube.[3] |

**Table 4** Search terms for boys between 5 and 8 years of age and evidence supporting each term

| Search term | Evidence supporting the search term |
|---|---|
| Challenge videos<br>SisVsBro challenges<br>Prank vlogs<br>Funny pranks<br>Cartoons<br>Let's play<br>Kids anime | Boys between 5 and 8 years who participated in study #1 and #4 watched challenge videos (e.g., SisVsBro), prank compilations and vlogs, animation (e.g., cartoons and anime), and Let's Play videos on YouTube.[1,4] |
| Minecraft gaming<br>Mario Kart gaming | Boys between 5 and 8 years who participated in study #1 and #5 watched Minecraft gaming videos.[1,2] In Canada, Minecraft and Mario Kart were the most popular video games among boys between 6 and 12 years in 2020.[5] |
| SpongeBob<br>StarWars Rebels<br>Miraculous | Boys between 5 and 8 years who participated in study #5 watched SpongeBob, Mickey Mouse, Peppa Pig, and Thomas the Tank Engine on YouTube.[7] While these were popular among children in the UK, in Canada, SpongeBob, StarWars Rebels, and Miraculous were the most popular television shows among children in 2023.[6-9]Thus, we replaced these search terms with the most popular television shows in Canada. |
| Paw Patrol unboxing<br>Batman unboxing<br>Lego unboxing | Children between 5 and 8 years who participated in study #1 reported watching toy unboxing videos.[1] Boys preferred to play with building and design toys, vehicles, and action figures.[1,10] In Canada, Paw Patrol toys, Batman toys, and Lego toys were the most popular among children for television, superhero, and building and construction toys, respectively, in 2021.[11] |

**Table 5** Search terms for girls between 5 and 8 years of age and evidence supporting each term

| **Search term** | **Evidence supporting the search term** |
|---|---|
| Playdoh creations<br>Fashion vloggers<br>Slime videos<br>Life hacks<br>How-to videos<br>How to make popsicles<br>Sophie Darcy<br>Mr. Kate<br>Rebecca Zomelo<br>SarahBeautyCorner | Girls between 5 and 8 years who participated in study #5 said that they enjoyed watching videos that taught them how to make things (e.g, popsicles), and videos featuring life hacks, playdoh creations, fashion vloggers (e.g, Sophie Darcy, and Rebecca Zomelo), slime videos, and vloggers such as SarahBeautyCorner and Mr. Kate.[2] |
| Polly Pocket unboxing<br>Barbie unboxing<br>Playmobil unboxing<br>Elsa and Anna unboxing<br>Winx dolls unboxing | Girls between 5 and 8 years who participated in study #1 and #5 reported watching toy unboxing videos (e.g., Polly Pocket, Barbie, Elsa and Anna, Playmobil, and Winx dolls).[1,2] Barbie was the most popular doll among children in Canada in 2021.[11] |

**Table 6** Search terms for boys between 2 and 4 years of age and evidence supporting each term

| **Search term** | **Evidence supporting the search term** |
|---|---|
| ABC song<br>Shapes song<br>Colours song<br>Days of the week song<br>Educational cartoons | Children between the age of 2 to 4 years who participated in study #3 watched educational cartoons and songs teaching shapes, colours, and early literacy skills, and music videos with 3D animation, all on YouTube.[12] |
| Sesame Street<br>Cocomelon | Children between the age of 2 to 4 years who participated in study #4 and #5 indicated that they watched Cocomelon and Sesame Street videos on YouTube.[2,4] In Canada, Sesame Street was one of the most popular shows among children in 2022.[13] Cocomelon ranked as one of the most popular shows among children in Canada in 2023.[14] |
| Baby Big Mouth Kids<br>Katie Cutie Kids TV<br>Little Angel & Friends<br>Busy Beavers<br>The Kiboomers | According to Social Blade, these channels are the top five made for kids YouTube channels among children in Canada.[15] All of these channels offer kids songs that are educational. |
| Hot Wheels play<br>Trains play<br>Action-figure play | Parents of children between 2 and 4 years who participated in study #1 and #4 indicated that their kids watched toy-playing videos (e.g., Hot Wheels play).[1,2] Boys also preferred to play with vehicles and action figures.[1,10] |

**Table 7** Search terms for girls between 2 and 4 years of age and evidence supporting each term

| Search term | Evidence supporting the search term |
|---|---|
| ABC song<br>Shapes song<br>Colours song<br>Days of the week song<br>Educational cartoons<br>Kids songs | Children between the age of 2 to 4 years who participated in study #3 watched educational cartoons and songs teaching shapes, colours, and early literacy skills, and music videos with 3D animations, all on YouTube.[12] |
| Sesame Street<br>Cocomelon | Children between the age of 2 to 4 years who participated in study #4 and #5 indicated that they watched Cocomelon, and Sesame Street.[2,4] In Canada, Sesame Street was one of the most popular shows among children in 2022.[13] Cocomelon ranked as one of the most popular shows among children in Canada in 2023.[14] |
| Baby Big Mouth Kids<br>Katie Cutie Kids TV<br>Little Angel & Friends<br>Busy Beavers<br>The Kiboomers | According to Social Blade, these channels are the top five made for kids YouTube channels among children in Canada.[15] All of these channels offer kids songs that are educational. |
| Dolls play<br>Teddy bear play | Parents of children between the age of 2 and 4 years who participated in study #4 and #1 indicated that their kids watched toy play videos.[1,2] Girls preferred to play with dolls and stuffed animals.[10] |

**Table 8** Search terms for women ≥18 years of age and evidence supporting each term

| Search term | Evidence supporting the search term |
|---|---|
| Healthy recipes<br>Healthy snacks<br>Yoga videos<br>Beauty tips<br>Skin care routine<br>Hair care routine<br>Self-care routine<br>Cuticle care<br>Fashion haul<br>Anti-aging supplements<br>Workout plan<br>Fashion trends<br>Home remedies<br>Mindfulness<br>Stress-management | Women who participated in study #8 reported watching health and wellness videos.[16]<br><br>According to Omnicore, beauty and style was the most watched content category on YouTube by women ≥18 years in the US in 2023.[17] |

**Table 9** Search terms for men ≥18 years of age and evidence supporting each term

| **Search term** | **Evidence supporting the search term** |
|---|---|
| Sports highlights<br>News today<br>Politics today<br>Technology news | Men who participated in study #8 reported watching technology-related videos, sports videos, and news videos.[16] |
| NHL highlights<br>NFL highlights<br>MLB highlights<br>CFL highlights<br>NBA highlights | Men who participated in study #8 reported watching sports videos and news on YouTube. The terms listed here were the top five most watched sports by adults ≥18 years in Canada in 2023.[18] |
| Gaming videos | According to Omnicore, gaming was one of the most watched content categories on YouTube by men in the US in 2023.[17] |
| Call of Duty<br>NHL 20 gameplay<br>Madden 20 gameplay<br>FIFA 20 gameplay<br>Grand Theft Auto | In Canada, these were the most popular online games played by men ≥18 years in 2020.[5] |

**Table 10** Marketing techniques and justification for including each as core child-targeted, broad child-targeted, or non-specific techniques

| Marketing technique | Justification |
|---|---|
| **Core child-targeted techniques** | |
| Presence of branded, licensed, and cartoon characters<br><br>Presence of celebrities and public figures | Studies have shown that spokes-characters including celebrities, branded, licensed and cartoon characters were used to target children on digital media platforms, product packaging, and television.[19-21] For instance, they were used to target children on their preferred websites in Canada (2018),[22] Facebook and YouTube food brand pages in New Zealand (2018),[23] and children's television programming in the US (2017).[24] These techniques have been shown to increase children's attention to advertisements, enhance product recognition, and lead to greater likability of the product.[25,26] |
| Child themes/visual design | This technique was used to target children on television, product packaging, and digital media platforms. [19-21] For example, it was used on children's preferred websites in Canada (2018),[22] Facebook and YouTube food brand pages in New Zealand (2018),[23] and children's television programming in the US (2017).[24] This technique has been shown to capture children's attention and influence their food preferences. Specifically, children rely on visual elements (animation, story format, and humour) when selecting their favourite food products and assessing their healthfulness.[27] |

| | |
|---|---|
| Use of child language | This technique was used to target children on television and digital media platforms. [19-21] For example, food marketers used this technique to target children on children's television programming in the US (2017)[24] and in Canada (2023).[28] This technique has been shown to attract children's attention and influence their food preferences.[20] |
| Appeals to fun/cool | This technique was used to target children across multiple platforms including television, digital media platforms, packaging, and print materials. [19-21] For instance, this technique was used to target children on YouTube videos of influencers popular with children in the UK (2019).[30] This technique has been shown to attract children's attention and influence their food choices.[31] |
| Advergames<br>Games and activities | Studies have shown that advergames and games and activities were used to target children on digital media platforms and product packaging. [19-21] For instance, they were used to target children on food company websites in the US (2017).[24] These techniques have been shown to negatively impact children's food attitudes and choices.[32] Advergames specifically blur the line between advertising and entertainment.[32,33] The marketing of unhealthy foods through advergames significantly increases children's consumption of unhealthy food.[34] |
| Unusual product flavour/shape | This technique was used to target children on digital media platforms including YouTube and television. [19-21] For instance, this technique was the technique that was most commonly used to target children on child-directed YouTube videos in Malaysia (2018).[35] This technique has been shown to influence children's attitudes and lead to increased consumption of marketed products.[36] |

| | |
|---|---|
| Presence of children<br>Adult-child situations | This technique was used to target children across multiple platforms including television, digital media platforms, packaging, and print materials. [19-21] For instance, child actors were used to target children on television in the US (2017)[24] and Canada (2023).[28] This technique may influence children's dietary patterns to align with those portrayed in the advertisement.[37] |
| **Broad child-targeted techniques** | |
| Appeals to health/nutrition<br>Displays of corporate and social responsibility<br>Promotion of product convenience<br>Other product claims<br>Price promotions, incentives, or giveaways<br>Cross-promotions<br>Appeals to flavour/texture | These marketing techniques were commonly used in previous literature to examine food marketing directed at children, despite not being typical child-targeted techniques. [19-21] These techniques may also target children's parents or caregivers, and therefore influence product purchases for children.[21,38] |
| **Non-specific techniques** | |
| Viral marketing<br>Calls-to-action<br>Advercation<br>Special effects<br>Songs/music | These techniques, when employed independently, were not deemed to be child-targeted since they may be employed to attract a broad audience, not exclusively children. Only when paired with core child-targeted techniques were they considered child-targeted. |

**Table 11** Artificial intelligence system performance measures

| Classification | Sensitivity (95% CI) | Specificity (95% CI) | Positive predictive value (95% CI) | Negative predictive value (95% CI) |
|---|---|---|---|---|
| **Food-related marketing instances** | 0.84 (0.77, 0.89) | 0.83 (0.79, 0.86) | 0.63 (0.56, 0.70) | 0.94 (0.90, 0.96) |
| **Unhealthy food marketing instances** | 0.85 (0.81, 0.89) | 0.58 (0.47, 0.70) | 0.89 (0.85, 0.92) | 0.51 (0.40, 0.61) |
| **Healthy food marketing instances** | 0.58 (0.47, 0.70) | 0.85 (0.81, 0.89) | 0.51 (0.40, 0.61) | 0.89 (0.85, 0.92) |
| **Core child-targeted techniques** | | | | |
| Branded characters | 0.64 (0.45, 0.80) | 0.91 (0.88) | 0.43 (0.29-0.58) | 0.96 (0.93 – 0.98) |
| Licensed characters | 1.00 (0.03, 1.00) | 1.00 (0.99-1.00) | 1.00 (0.03-1.00) | 1.00 (0.99-1.00) |
| Celebrities / public figures | 0.76 (0.50, 0.93) | 0.99 (0.97, 1.00) | 0.72 (0.47, 0.90) | 0.99 (0.97, 1.00) |
| Other cartoon characters | 0.27 (0.08, 0.55) | 0.99 (0.97, 1.00) | 0.50 (0.16, 0.84) | 0.97 (0.94, 0.98) |
| Child themes/visual design | 1.00 (0.97, 1.00) | 0.00 (0.00, 0.02) | 0.54 (0.49, 0.59) | 0.00 (0.00, 0.97) |
| Use of child language | 0.19 (0.12, 0.27) | 0.84 (0.79, 0.88) | 0.34 (0.23, 0.47) | 0.69 (0.64, 0.74) |
| Appeals to fun/cool | 0.88 (0.82, 0.93) | 0.11 (0.07, 0.16) | 0.47 (0.41, 0.52) | 0.51 (0.35, 0.67) |
| Games and activities | 0.04 (0.00, 0.18) | 0.99 (0.98, 1.00) | 0.25 (0.01, 0.81) | 0.93 (0.90, 0.95) |
| Unusual product flavour | 0.12 (0.01, 0.36) | 0.97 (0.95, 0.99) | 0.18 (0.02, 0.52) | 0.96 (0.93, 0.98) |
| Unusual product shape | 0.80 (0.59, 0.93) | 0.37 (0.32, 0.42) | 0.08 (0.05, 0.12) | 0.96 (0.92, 0.99) |
| Adult-child situations | 0.96 (0.85, 0.99) | 0.26 (0.21, 0.31) | 0.15 (0.11, 0.20) | 0.98 (0.92, 1.00) |
| **Broad child-targeted techniques** | | | | |

| | | | | |
|---|---|---|---|---|
| Appeals to health/nutrition | 0.48 (0.41, 0.56) | 0.56 (0.49, 0.63) | 0.50 (0.42, 0.57) | 0.55 (0.48, 0.62) |
| Displays of corporate and social responsibility | 0.02 (0.00, 0.09) | 0.99 (0.97, 1.00) | 0.25 (0.01, 0.81) | 0.91 (0.87, 0.94) |
| Promotion of product convenience | 0.19 (0.09, 0.31) | 0.86 (0.82, 0.90) | 0.19 (0.09, 0.31) | 0.86 (0.82, 0.90) |
| Price promotions, incentives, or giveaways | 0.36 (0.24, 0.49) | 0.72 (0.66, 0.77) | 0.20 (0.13, 0.29) | 0.85 (0.80, 0.89) |
| Cross-promotions | 0.19 (0.05, 0.42) | 0.81 (0.77, 0.85) | 0.06 (0.02, 0.14) | 0.94 (0.91, 0.97) |
| Appeals to flavour/texture | 0.58 (0.51, 0.64) | 0.56 (0.47, 0.65) | 0.72 (0.65, 0.78) | 0.41 (0.34, 0.49) |
| **Non-specific techniques** | | | | |
| Viral marketing | 0.00 (0.00, 0.46) | 0.99 (0.96, 1.00) | 0.00 (0.00, 0.84) | 0.98 (0.98, 0.99) |
| Calls-to-action | 1.00 (0.97, 1.00) | 1.00 (0.99, 1.00) | 1.00 (0.97, 1.00) | 1.00 (0.99, 1.00) |
| Special effects | 0.43 (0.37, 0.47) | 0.60 (0.26, 0.88) | 0.97 (0.94, 0.99) | 0.03 (0.01, 0.06) |
| Songs/music | 0.98 (0.93, 1.00) | 0.33 (0.04, 0.78) | 0.99 (0.95, 1.00) | 0.12 (0.01, 0.36) |

CI, confidence interval.
Analyses were conducted using R with manual labels serving as true positives.

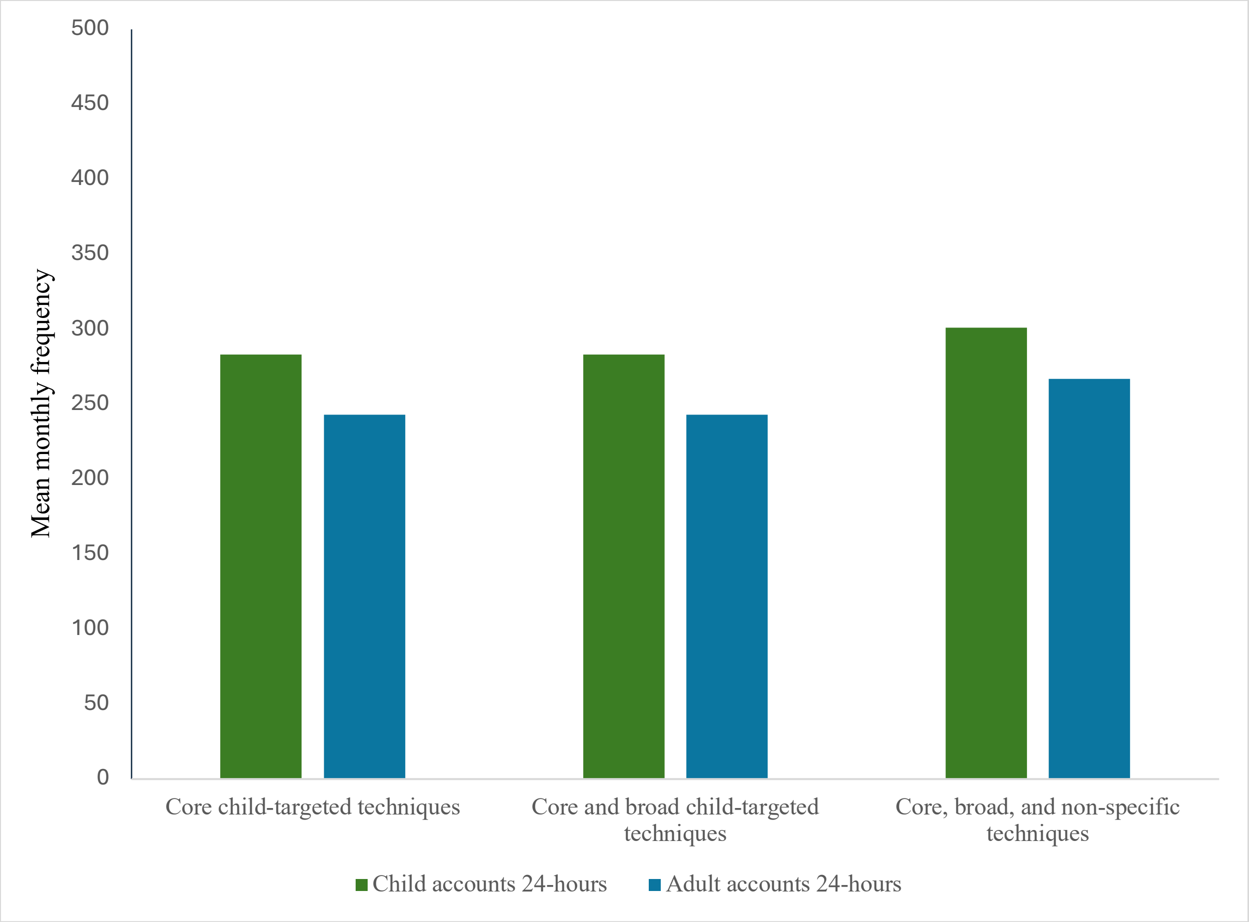


**Figure. 1** The mean monthly frequency of unhealthy food marketing that was child-targeted shown to all child and adult accounts based on three different definitions of child-targeted food marketing[a,b]. Analyses were conducted using Friedman tests. For both child and adult accounts, no significant differences were found in the mean monthly frequency of child-targeted unhealthy food marketing instances when using the three different definitions of child-targeted food marketing.

[a] Mean monthly frequency for the four child and two adult accounts that were shown food marketing
[b] No food marketing instances were shown to the child accounts 2-4 years.

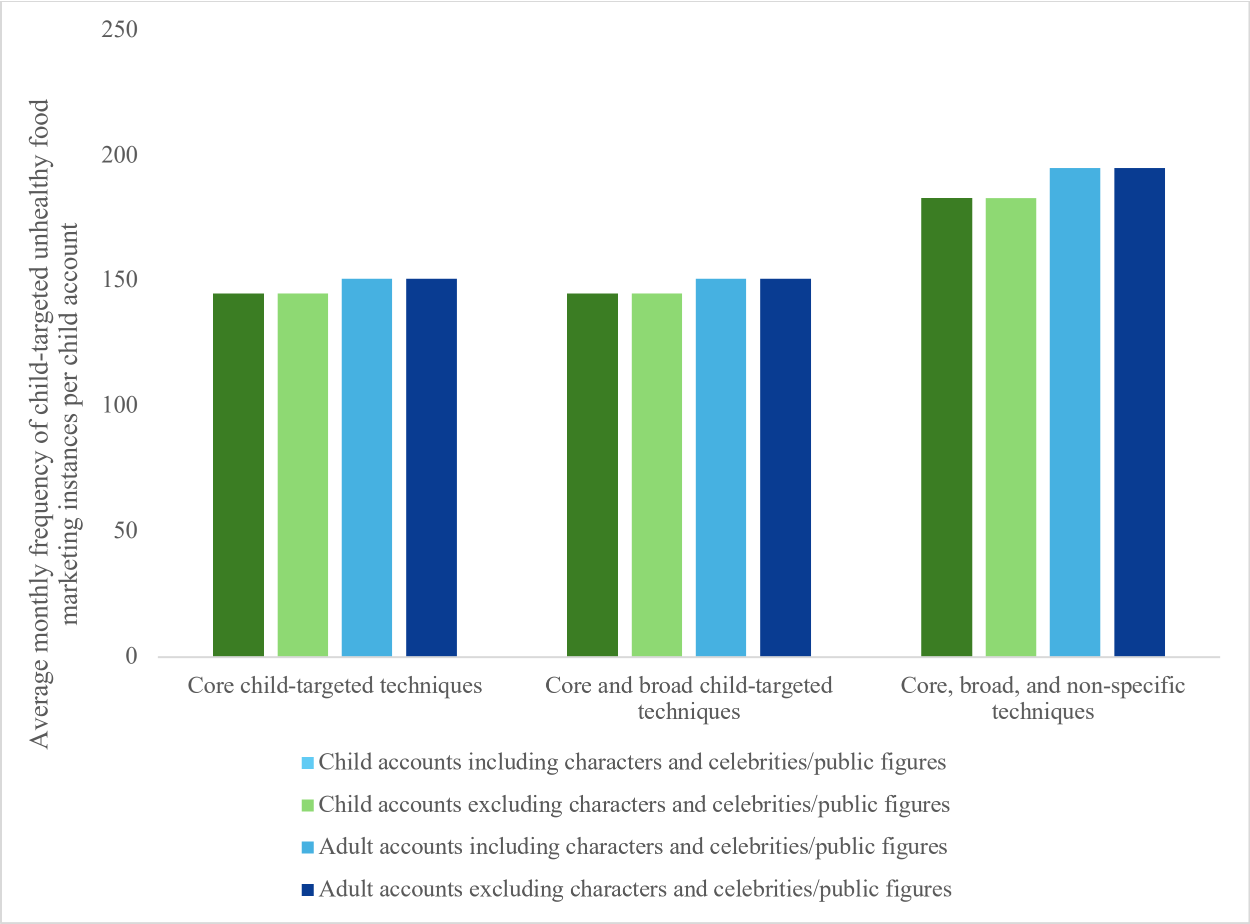


**Figure. 2** The mean monthly frequency of child-targeted unhealthy food marketing instances shown to all child and adult accounts based on three different definitions of child-targeted food marketing when including and excluding the presence of presence of characters and celebrities/public figures.[a,b,c] Analyses were conducted using Friedman tests. For both child and adult accounts, no significant differences were found in the mean monthly frequency of child-targeted unhealthy food marketing instances when using the three different definitions of child-targeted food marketing when including characters and celebrities/public figures. Additionally, there were no significant differences in the frequency of child-targeted marketing instances within each definition when including or excluding the presence of characters and celebrities/public figures.

[a] Mean monthly frequency for the four child and two adult accounts that were shown food marketing [b] No food marketing instances were shown to the child accounts 2-4 years.

[c] This analysis was performed on a smaller subset of the available data (n= 2,599 food marketing instances) as some marketing instances were no longer available on YouTube at the time of this analysis.

**Table 12** The nature of child-targeted unhealthy food marketing instances shown to child and adult accounts by age and gender when including characters and celebrities/public figures

| **Marketing Technique** | **Mean monthly frequency (%) based on a 24-hour or a 2-hour daily viewing period** | | | | | | | | | | | |
|---|---|---|---|---|---|---|---|---|---|---|---|---|
| | **Boy 5-8 years** | | **Girl 5-8 years** | | **Boy 9-12 years** | | **Girl 9-12 years** | | **Man ≥ 18 years** | | **Woman ≥ 18 years** | |
| | **24-hours** | **2-hours** | **24-hours** | **2-hours** | **24-hours** | **2-hours** | **24-hours** | **2-hours** | **24-hours** | **2-hours** | **24-hours** | **2-hours** |
| Presence of branded characters | 10 (9.3) | 1 (11.1) | 14 (8.6) | 1 (7.1) | 12 (8.0) | 1 (8.3) | 14 (7.3) | 1 (6.3) | 6 (4.7) | 0 (0.0) | 16 (8.8) | 1 (6.7) |
| Presence of other cartoon characters | 4 (3.7) | 0 (0.0) | 11 (6.8) | 1 (7.1) | 10 (6.7) | 1 (8.3) | 13 (6.7) | 1 (6.3) | 6 (4.7) | 0 (0.0) | 14 (7.7) | 1 (6.7) |
| Presence of licensed characters | 1 (0.9) | 0 (0.0) | 7 (4.3) | 0 (0.0) | 2 (1.3) | 0 (0.0) | 7 (3.6) | 0 (0.0) | 1 (0.8) | 0 (0.0) | 1 (0.5) | 0 (0.0) |
| Presence of celebrities/ public figures | 4 (3.7) | 0 (0.0) | 8 (4.9) | 0 (0.0) | 5 (3.3) | 0 (0.0) | 8 (4.1) | 0 (0.0) | 8 (6.2) | 0 (0.0) | 10 (5.5) | 1 (6.7) |

Analyses were conducted using negative binomial Poisson regression followed by pairwise comparisons of estimated marginal means to examine differences in frequency by age and gender.
The denominator for calculating the proportion of each listed technique is the total monthly frequency of child-targeted unhealthy food marketing instances shown to each account that used any core child-targeted technique.
The total viewing time in a month per account was 672 hours (24 hours x 7 days x 4 weeks) for a continuous 24-hour daily viewing period and 56 hours (2 hours x 7 days x 4 weeks) for a 2-hour daily viewing period.
No food marketing instances were shown to the child accounts 2-4 years.
This analysis was conducted on a subsample of 2,599 of food marketing instances as some marketing instances were no longer available on YouTube at the time of this analysis.

*References*

and-streaming-market-share-analysis-2022-netflix-nbc-cbs-abc-and-hbo/. Accessed 30 Jan 2025.

14. Cocomelon (YouTube): Canada daily TV audience insights for smarter content decisions - Parrot Analytics. https://tv.parrotanalytics.com/CA/cocomelon-youtube. Accessed 10 Oct 2023.
15. Socialblade.com. https://socialblade.com/youtube/top/country/ca. Accessed 9 Oct 2023.
16. Landrum AR. Are women a missing audience for science on YouTube? An exploratory study. Front Commun. 2021;6:610920.
17. Omnicore Agency. YouTube by the numbers (2024): Stats, demographics & fun facts. In: Omnicore Agency. 2024. https://www.omnicoreagency.com/youtube-statistics/. Accessed 9 Oct 2024.
18. Angus Reid Institute. Super-ior Bowl? Most Canadian football fans would choose Super Bowl over Grey Cup, but not everywhere. In: Angus Reid Institute. 2023. https://angusreid.org/wp-content/uploads/2023/06/2023.06.20_LIV_Merger_tables.pdf. Accessed 17 Oct 2023.
19. World Health Organization. Food marketing exposure and power and their associations with food-related attitudes, beliefs and behaviours: a narrative review. Geneva: WHO; 2022. Contract No.: CC BY-NC-SA 3.0 IGO.
20. Elliott C, Truman E. Measuring the Power of Food Marketing to Children: A Review of Recent Literature. Curr Nutr Rep. 2019 Dec; 8(4): 323-332.
21. Mulligan, C., Potvin Kent, M., Christoforou, A.K. et al. Inventory of marketing techniques used in child-appealing food and beverage research: a rapid review. Int J Public Health 65, 1045–1055 (2020).
22. Potvin Kent, M., & Pauzé, E. (2018). The effectiveness of self-regulation in limiting the advertising of unhealthy foods and beverages on children's preferred websites in Canada. Public Health Nutrition, 21(9), 1608-1617.
23. Vandevijvere S, Aitken C, Swinburn B. Volume, nature and potential impact of advertisements on Facebook and YouTube by food brands popular in New Zealand. N Z Med J. 2018; 131(1473):14–24
24. Vilaro MJ, Barnett TE, Watson AM, Merten JW, Mathews AE. Weekday and weekend food advertising varies on children's television in the USA but persuasive techniques and unhealthy items still dominate. Public Health. 2017;142:22–30.
25. Mizerski R. The Relationship between Cartoon Trade Character Recognition and Attitude toward Product Category in Young Children. J Marketing. 1995; 4: 58-70.
26. Neeley SM, Sherman DW. Using animated spokes-characters in advertising to young children. J Advertising 2004; 3: 7–23.
27. Letona, P., Chacon, V., Roberto, C. et al. A qualitative study of children's snack food packaging perceptions and preferences. BMC Public Health 14, 1274 (2014).
28. Potvin Kent M, Soares Guimaraes J, Pritchard M, Remedios L, Pauzé E, L'Abbé M, Mulligan C, Vergeer L, Weippert M. Differences in child and adolescent exposure to unhealthy food and beverage advertising on television in a self-regulatory environment. BMC Public Health. 2023 Mar 23;23(1):555.
29. LoDolce ME, Harris JL, Schwartz MB. Sugar as part of a balanced breakfast? What cereal advertisements teach children about healthy eating. J Health Commun. 2013;18(11):1293-309.
